# Cross-validation coordinate analysis (CVCA): performing coordinate based meta-analysis using cross-validation

**DOI:** 10.1101/2024.03.13.24304212

**Authors:** C.R. Tench

## Abstract

Coordinate based meta-analysis (CBMA) can be used to estimate where a future neuroimaging study testing a particular hypothesis might report summary results (activation foci, for example). However, current methods cannot be validated for all possible analyses, and use empirical features that might not always be ideal. Indeed, the various algorithms that perform CBMA tend to produce somewhat different results even on the same data. Furthermore, the use of null hypothesis significance testing (NHST) in the algorithms is not strictly in keeping with meta-analysis, where the aim is usually effect estimation rather than statistical significance.

Given the limitations of current CBMA algorithms, a new algorithm has been developed that will perform its analysis using cross validation: cross-validation coordinate analysis (CVCA). Although a full validation cannot be performed, CVCA optimises its parameters in such a way as to make the analysis results most similar to the held-out data. The algorithm can be used as a stand-alone meta-analysis method, or to provide confidence in results from other algorithms where no validation is performed.

Software to perform CVCA is freely available.

## Introduction

### What is CBMA

Coordinate based meta-analysis (CBMA) is commonly used to summarise effects reported by multiple independent, but related by a shared hypothesis, neuroimaging studies. It is employed to meta-analyse (amongst others) voxel-based morphometry (VBM) or functional magnetic resonance imaging (fMRI) and uses only reported summary results such as coordinates (foci of activation, for example). By analysing multiple studies simultaneously, those results that represent a common finding among at least some of the studies are identified and are assumed indictive of relevant and reliably measurable effects. In the absence of whole brain statistical images with which to perform full image based meta-analysis (IBMA), CBMA can provide a useful summary of where studies testing the same hypothesis are likely to report significant results.

In meta-analysis the aim is to obtain better estimates of an effect that has been measured independently multiple times. Effects considered are typically reported with an associated variance, and the meta-analysis uses both. In CBMA the effects are the reported foci (although this can be augmented with further detail such as Z scores or t statistics), but there is no accompanying spatial variance estimate. This leaves multiple independent effects reported per study without any clear way to relate those reported in one study to those reported in other studies. In CBMA an empirical estimate of the unknown spatial variance is used to associate the coordinates reported by different studies. This, along with null hypothesis significance testing (NHST), is used to cluster the independent findings into an estimated meta-effect.

Probably the most popular method of performing CBMA is the activation likelihood estimate (ALE) algorithm (Turkeltaub et al., 2002). Smoothing the reported foci using a Gaussian kernel accounts for spatial variance of activation peaks. A modelled activation image for each independent study results from application of the kernel to each focus reported by the study, and these are subsequently combined into a single ALE map representing the likelihood of activation. A permutation test, based on randomising foci uniformly in a specified image space, is performed to establish a statistical threshold with the aim of identifying regions of the brain reported with high concordance in comparison to just random chance. Post threshold, voxels with statistically significant ALE provide an estimate of the distribution of reported foci relating to the common hypothesis. It is the anatomical location of the peaks in this distribution that form the output of the ALE algorithm and the result on which the interpretation and conclusion are based.

Another popular approach is the kernel density analysis (KDA) (Wager et al., 2007) method, which aims to estimate the density of reported activation foci. Rather than the probabilistic model employed by ALE to reflect likelihood of activation, KDA uses the kernel density method commonly used for probability density estimation (Epanechnikov, 1969).

Besides these algorithms there are multiple others that have similar aims but using different specific assumptions to perform the analysis. Amongst these algorithms there are a range of null hypotheses, methods of statistical thresholding, and type and size of kernel. For example, most algorithms employ randomisation of coordinates into an image space to represent an empirical null hypothesis reflecting the situation of no systematic spatial agreement across studies. However, the signed differential mapping permutation of subject images (SDM-PSI) method (Albajes-Eizagirre et al., 2018) and the parametric coordinate based meta-analysis (PCM) method (Costafreda, 2012) make distributional assumptions about the reported statistical effects and employ a null of zero effect (reported Z scores or t statistics), and consequently have an extra study inclusion criteria that both positive and negative effects (activation & deactivation, for example) are reported in an unbiassed way; studies performing one tailed analyses would violate the assumption of these methods and cannot be included. These differences can produce a range of results from the same data (Ferreira and Busatto, 2010). The conclusions of studies performing CBMA may therefore depend on the algorithm utilised, and one sensible option might be to perform the analysis with multiple different methods to obtain an overview.

There have been attempts to compare CBMA methods in a validation setting, whereby the results according to various algorithms are directly compared to a relevant standard. However, validation of CBMA is problematic since the it is only strictly relevant to the specific study used as the ground truth and the set of studies being meta-analysed. One attempt at validating various CBMA methods uses a ground truth provided by IBMA (Salimi-Khorshidi et al., 2009); IBMA is preferred to CBMA if the data are available. The study attempted to choose CBMA kernels that would maximise the similarity to IBMA results. The authors point out that while optimal kernels can be found, this is only valid for the specific data being considered. An example of validating thresholds is provided in (Radua and Mataix-Cols, 2009), where simulation (unreported) analysis supports an uncorrected p-value threshold of 0.001 for the signed differential mapping (SDM) method. In a later evolution of the method this was increased to 0.005. In neither case would unprincipled thresholding be valid for studies exhibiting no meta-effect, which has previously lead to some contrary results (Rahimi-Jafari et al., 2022; Tanasescu et al., 2015; Zhong et al., 2016). Another study has validated the width of the Gaussian kernel used in the ALE algorithm, concluding that a fixed standard deviation of 8.4mm is optimal, with an additional width representing an increasing uncertainty with decreasing number of subjects within study (Eickhoff et al., 2009). However, the procedure used to estimate the width did not include any random effect. Furthermore, using a fixed standard deviation with respect to the number of studies in the analysis produces results that do not converge as the number of studies contributing increases, causing the significant clusters of voxels to expand rather than reaching a stable volume (Belyk et al., 2017; Eickhoff et al., 2016; Frahm et al., 2023; Tench et al., 2014). Indeed when estimating a density using kernels the kernel width should be reducing with sample size (Epanechnikov, 1969), which for CBMA is the number of studies.

In general, the various methods of thresholding and the kernels used in CBMA cannot be validated in a way that applies to all possible uses of the algorithms. As such, these need to be implemented pragmatically, and the limitations accepted. However, it is possible perform CBMA using cross-validation. Here, leave one out (LOO) cross-validation is employed to inform kernel widths and thresholds that constrain the results of N analyses (N = total number of studies being analysed) conducted on N-1 of the studies to be most similar to, on average, the results of the 1 held-out study. Unlike other algorithms, the results do not rely only on an empirical kernel width and a threshold that considers only the type 1 error rate. The approach detailed here is referred to as cross-validation coordinate analysis (CVCA). Software to perform CVCA is freely available https://www.nottingham.ac.uk/research/groups/clinicalneurology/neuroi.aspx.

## Methods

### Cross-validation using leave one out

Briefly, the CVCA approach produces an estimation of the density of reported foci using a kernel density estimation method. Two parameters determine the results: kernel width (*ω*), and a minimum number of studies (*N*_*min*_) defining regions of high concordance among the studies. The algorithm makes use of the fact that the meta-results are identified as areas of high density (high concordance) of foci reported by independent studies, which are distinct from results that are study specific where the reported foci are lower density. In 3 dimensions the smallest value considered for *N*_*min*_ is 4, because a 3D mean and at least a spatial variance are needed to describe the local density being estimated. For the given *N*_*min*_ and total number of studies N, the width of the kernel is optimised by obtaining the meta-results using N-1 studies, then maximising a density score, which reflects how closely foci from 1 held-out study agree with the meta-results on average (over the different held-out studies). The idea is that the score initially increases as the meta-results form with increasing kernel width; consider that a kernel width of zero is likely to produce no meta-results. Once the meta-results have formed, increasing the width further causes the kernel to ‘spill’ out beyond the meta-results into the background, and this is used to cause a reduction in the score. In this way the score has a peak that is considered the optimal solution.

### Kernel density method

This algorithm uses a kernel to define a density reflecting the number of independent studies reporting effects in a similar location, which is the meta-effect of interest. The aim is to use cross-validation to choose parameters to maximise the mean density at the reported foci from the left-out study. In this approach focus *i* in study *j* at location ***f***_*ij*_ has a density at location ***r*** of

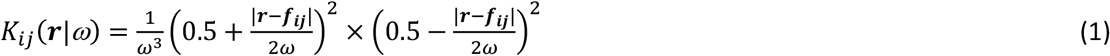

given the kernel width *ω*; note that *ω*^*3*^ normalises the kernel density (except for an unimportant multiplicative constant), and the kernel is assigned a value of zero if |***r***-***f***_*ij*_|≥*ω*. The kernel is based on a beta distribution parameterised such that it appears bell shaped and somewhat like the Gaussian kernel employed in some CBMA algorithms but having only limited extent such that very distal foci cannot influence the density estimate. This is depicted in figure 1.

**Figure 1.**
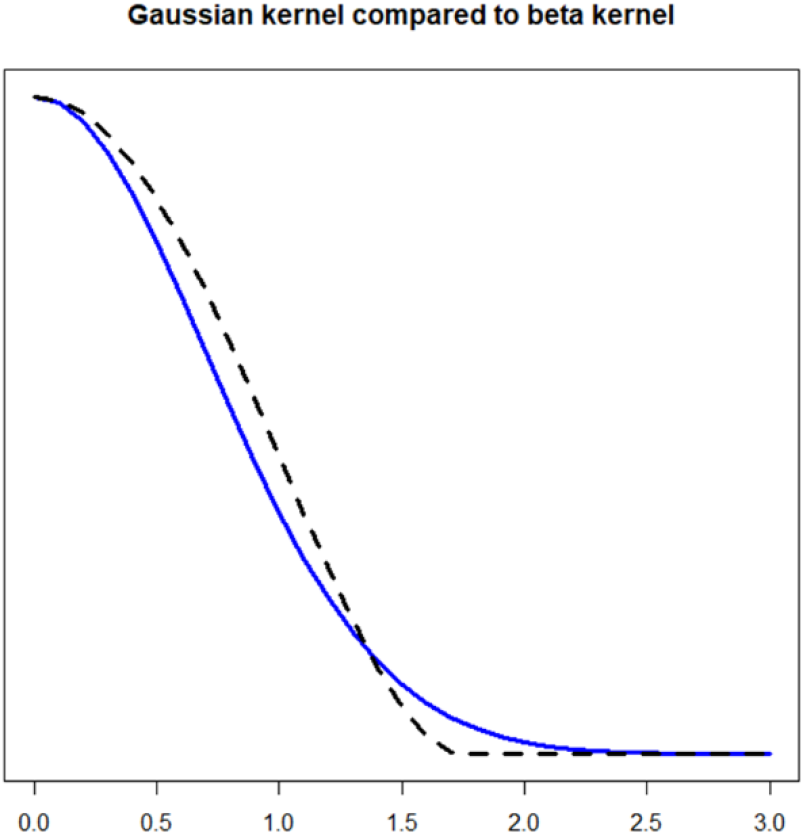
The standard Gaussian kernel profile (blue line) compared to the beta distribution based kernel profile used in CVCA (dashed black line). The beta kernel reaches zero at width ∼1.75, so is similar to a Gaussian kernel with standard deviation of *ω*/1.75.

Other than the kernel width parameter there is also parameter *N*_*min*_. This parameter defines *concordant foci*: foci that are located within distance *ω* of at least *N*_*min*_ -1 foci from *N*_*min*_*-*1 other studies in the set of studies *S*. These foci are suggestive of a local meta-effect, since the effect is reported independently by multiple studies. An indicator variable for the *i*^th^ focus in study *j* (*I*_*ij*_(*S*)) has a value of 1 if the focus is concordant and zero otherwise.

The cross-validation analysis incorporates these definitions to create a density score that reflects how well the left-out study matches the meta-analysis results from the other studies. Let the set of studies *S*_*-k*_ be the set of all studies except left-out study *k*. Then, given parameters *ω* and *N*_*min*_, the total score is

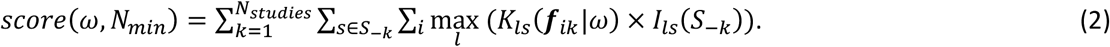

In words this score is a sum of the kernel density from the closest concordant foci in each kept-in study to each focus in the left-out study, summed over all leave-out studies.

One issue with the score described by equation (2) is that concordant foci may contribute multiple times within a single left-out study if the reported foci from that study fall close together. This can occur, for example, if multiple experiments performed on the same subjects are included in the analysis grouped by study, as suggested in (Turkeltaub et al., 2012). This has been considered in CVCA, for each left-out study, by noting which concordant foci appear in the sum (equation (2)) multiple times and then adding only the average contribution rather than the sum.

In CVCA the chosen kernel width maximises the score function (equation (2)) for a given *N*_*min*_. The score is increasing with *ω* when the width is small, because wider kernels produce more concordant foci to contribute to the sum. The purpose of the *ω*^*-3*^ normaliser in equation (1) is to cause the score to reduce as the kernel width extends beyond the concordant regions. Optimisation of kernel width is conducted for each value of *N*_*min*_, with the chosen solution producing the largest maximum score overall.

### Null hypothesis

CVCA results do not make use of statistical significance, which is in keeping with meta-analysis as a way to estimate effect. The algorithm works only by adjusting the kernel widths and a threshold in a way that is not based on p-values. Nevertheless, it is important that the results score highly at being similar to the held-out studies on average compared to the when the studies do not suggest any meta-effect. A null hypothesis can be derived using random foci, as used by the popular ALE algorithm. A single p-value, which is expressed as a Z score, is then produced comparing the score from the observed data with scores from the random foci. Since this is a single p-value, there is no requirement for controlling the familywise error rate, for example.

### Organising studies by subject group

When conducting CBMA the foci from multiple studies must be extracted. However, within study it is common that multiple experiments are performed on the same subjects, often with similar hypotheses. The different within-study-group experiments are not independent and cannot be considered so in the meta-analysis without causing bias. A proposed solution is to arrange all foci by study, rather than experiment (Turkeltaub et al., 2012). However, a consequence for methods that spatially randomise foci independently to generate the null distribution, such as the popular ALE algorithm, is a reduction in the sensitivity to meta-effects due to the number of foci randomised being larger than the number of independent effects (Tench et al., 2022). The solution suggested here is to include only one experiment from each study, and then use the other experiments to perform sensitivity analysis to make sure results are robust to perturbation.

## Experiments

### Thermal pain in HC

The first experiment demonstrates the optimisation of the kernel width using reported fMRI summary data from painful stimulus studies in healthy volunteers. The data have been utilised previously, and are made available http://doi.org/10.17639/nott.7095. This data are also used to demonstrate how the kernel width changes as smaller subsets of the studies are analysed, and compared to other CBMA algorithms: ALE and Analysis of Brain Coordinates (ABC) (Tench et al., 2022).

### AD and face discrimination

As further examples of how CVCA performs compared to ALE, data collected for a recent ALE methods study (Costa et al., 2023) are used. The available data (Costa et al., 2023) are not organised by subject group, so to avoid bias they are modified to include only a single experiment per author; the first occurrence of each author in the file. The first example is a meta-analysis of structural Alzheimer’s disease studies, and the second a meta-analysis of face discrimination fMRI task studies.

### Narcolepsy

As a demonstration of CVCA results with data that have been found to not demonstrate any meta-effect (Rahimi-Jafari et al., 2022; Tanasescu et al., 2015). Data published previously (Rahimi-Jafari et al., 2022) are used, representing neuroimaging results of narcolepsy.

## Results

### Painful stimulus studies

Eighty-three independent fMRI studies of painful stimulus of healthy controls were analysed. Figure 2 shows how the meta-results change as the kernel width is altered. The score (figure 2, bottom right) peaks when the kernel width is 7mm, and the parameter *N*_*min*_ is 4. From the figure the features of the CVCA approach are apparent from the spatial extent of the kernel density (shown as a light grey background to the colour foci) compared to the foci forming that density. When the kernel is too narrow, the density does not extend sufficiently far to include all densely packed foci, whereas when the kernel is too wide the density extends beyond the foci, reducing the score thanks to the normalisation in equation (1).

**Figure 2.**
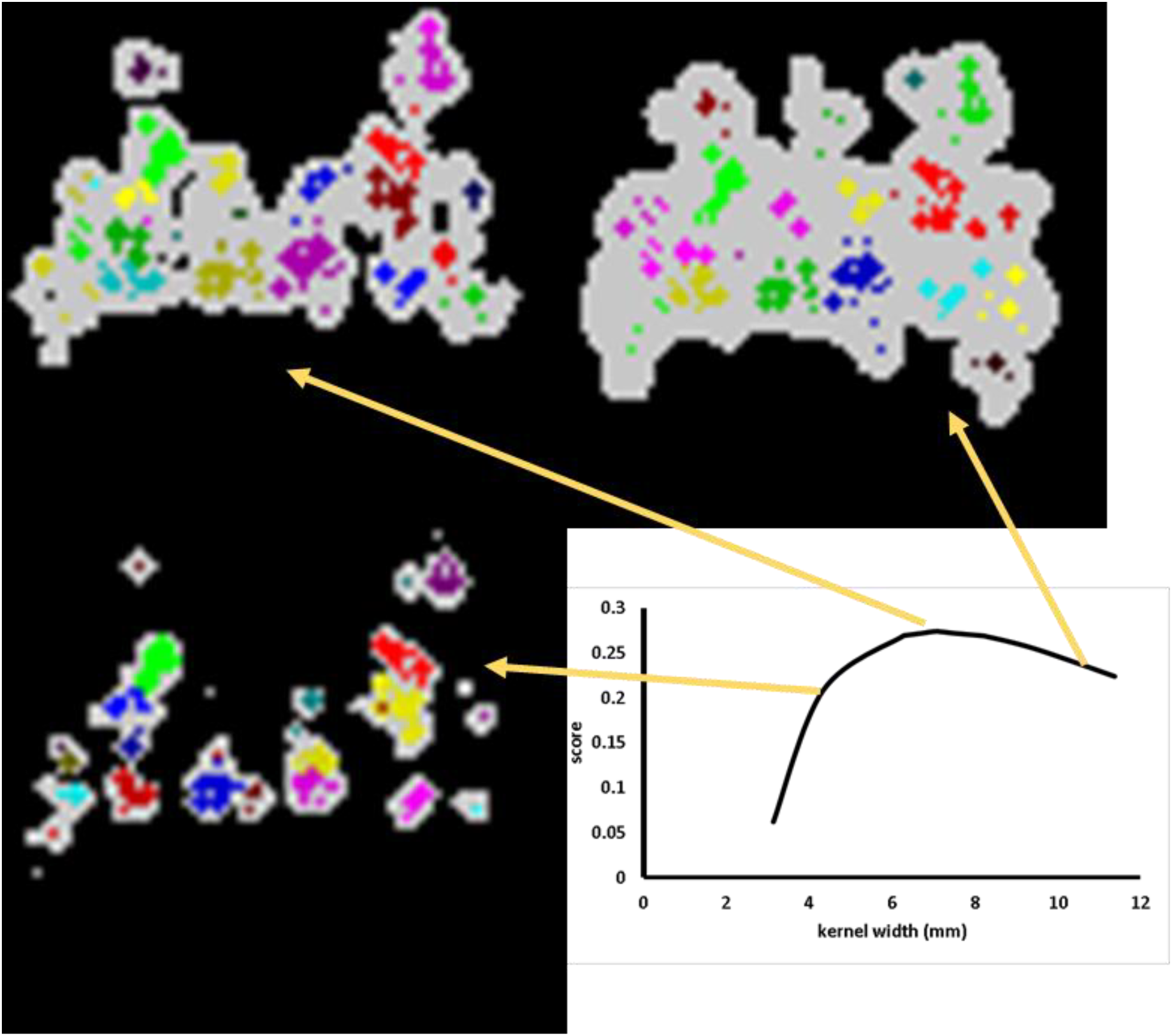
The optimisation of kernel width for the thermal pain study. When the kernel width is 5mm, the regions of high study density (light grey pixels) are small, being formed by few concordant foci (coloured markers). At the peak score (kernel width 7mm), the regions of high study density have grown and include more concordant foci. At a kernel width of 11mm, the high-density regions have expanded by comparison to those at a kernel width of 7mm, but the concordant foci have not increased to fill the density. This behaviour is reflected in the score, which is modulated firstly by the formation of high study density regions, which increases the score, then by the over extension of the high study density regions beyond the concordant foci, which reduces the score by the action of the normalisation of the kernel.

The CVCA results on the painful stimulus data are shown in figure 3; for all 83 studies as well as for subsets of 30 and 10 studies. Also shown, for comparison, are the results from the ALE and ABC algorithms. Both the ALE and ABC algorithms employ NHST, which produces a reducing amount of significant effect as the number of studies reduces. The impact of reducing the number of studies appears to have a lesser impact on the ABC algorithm compared to the ALE algorithm in this case, which may be due the fixed kernel width in the ALE algorithm; ABC has an adaptive kernel width that can adjust to the number of studies. By contrast this figure shows a different relationship between number of studies and apparent detail in the density images from the CVCA algorithm. As might be expected for a meta-analysis, increasing the number of studies analysed increases detail in the estimate of reported foci density.

**Figure 3.**
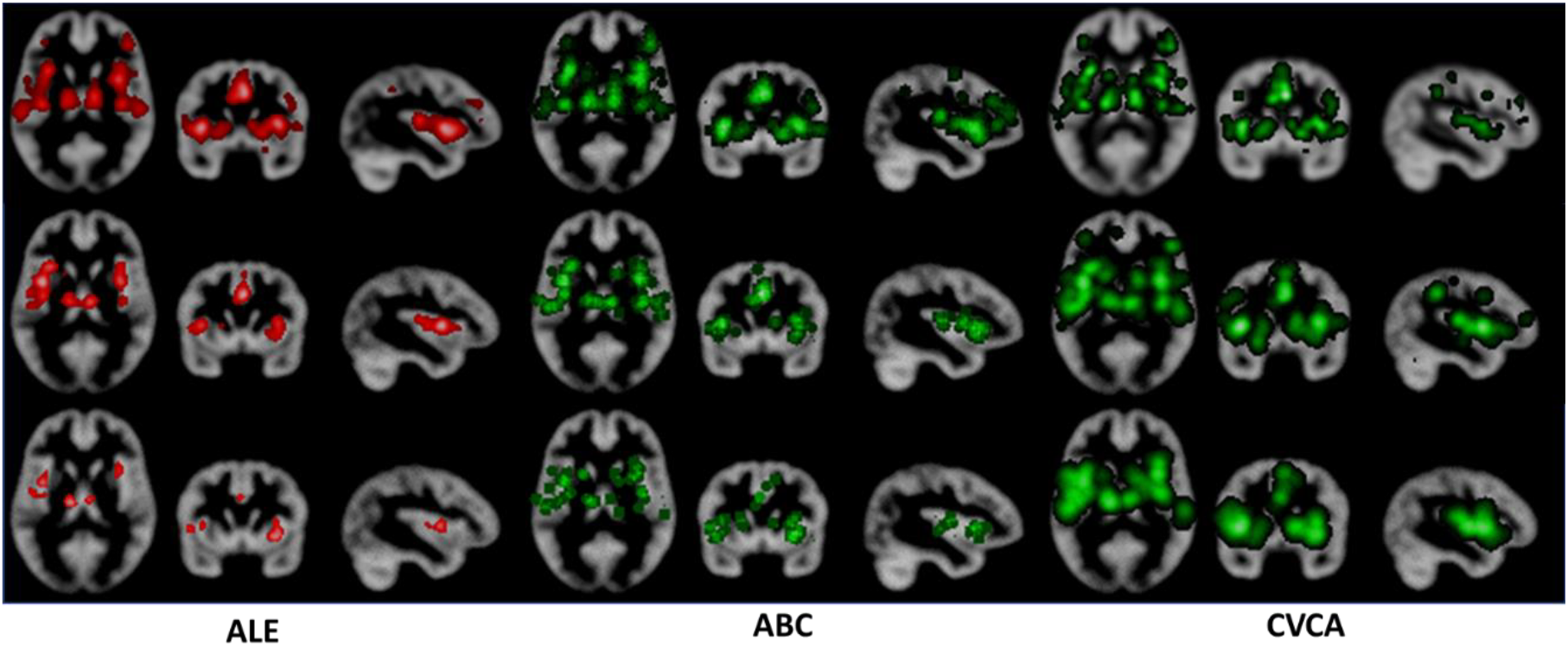
Example results using ALE, ABC and CVCA for the thermal pain experiment. The top figure shows results for all 83 studies, middle is for the first 30 (in the text file) studies, and bottom shows results for the first 10 studies. In CVCA the kernel widths were increasing with decreasing number of studies: 7mm, 10.6mm and 13.5mm respectively. There are fewer significant results from ALE and ABC when the number of studies analysed is lower, as expected for null hypothesis significance testing. Contrast this with the results from CVCA, where there is reducing detail (increasing image blurring) with fewer studies analysed.

Figure 4 shows the CVCA result of null hypothesis significance testing using random foci to generate the null. The score is very significantly higher than the null samples, expressed as a Z score of 48. The Z score is computed using just 20 random samples; more would be required to estimate this accurately, but since the Z score is so large any inaccuracy would be unlikely to change the conclusion.

**Figure 4.**
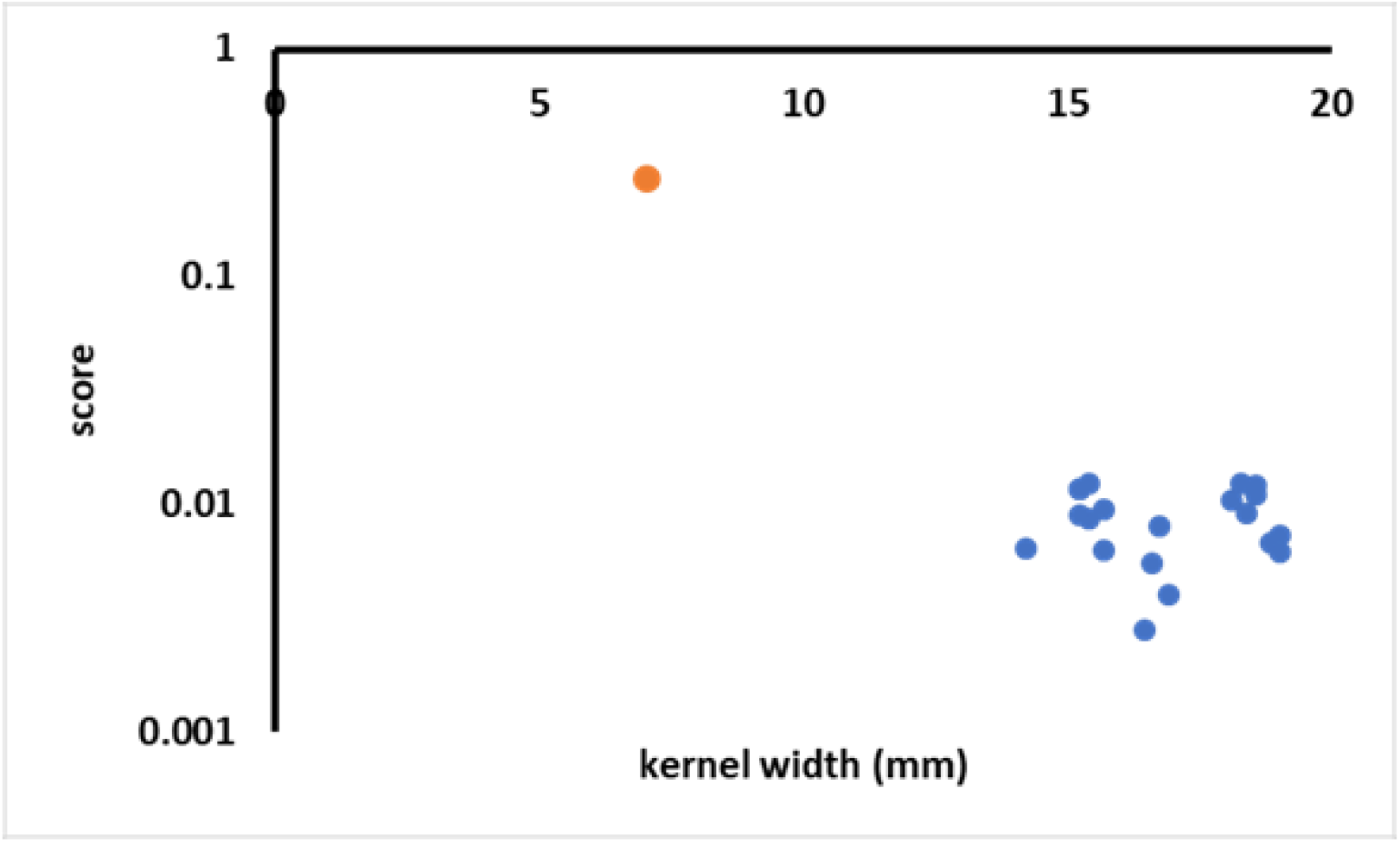
This shows, for the 83 thermal pain studies, a scatter plot of score and kernel width for the reported data (orange circle) and data randomly distributed and having null meta-effect (blue circle). The reported data has significantly higher score, and smaller kernel width, than the null data. If the reported data point fell within the cloud of null data, the results would not be significant (would not be distinct from the null). In this example, only 20 null samples are used since the distinction in score from the null is clear with a Z score of 48.

### Alzheimer’s disease and face discrimination tasks

The Alzheimer’s disease analysis involved 35 structural studies, and results are shown in figure 5 along with the analysis from the ALE algorithm. The two methods produce similar results in this example. The face discrimination analysis involved 18 functional studies, and again both methods produced similar results. The CVCA parameters were: kernel width=5.5mm and *N*_*min*_ was 4 for the Alzheimer’s analysis, and for face discrimination the kernel width was 8.5mm and *N*_*min*_ was 4.

**Figure 5.**
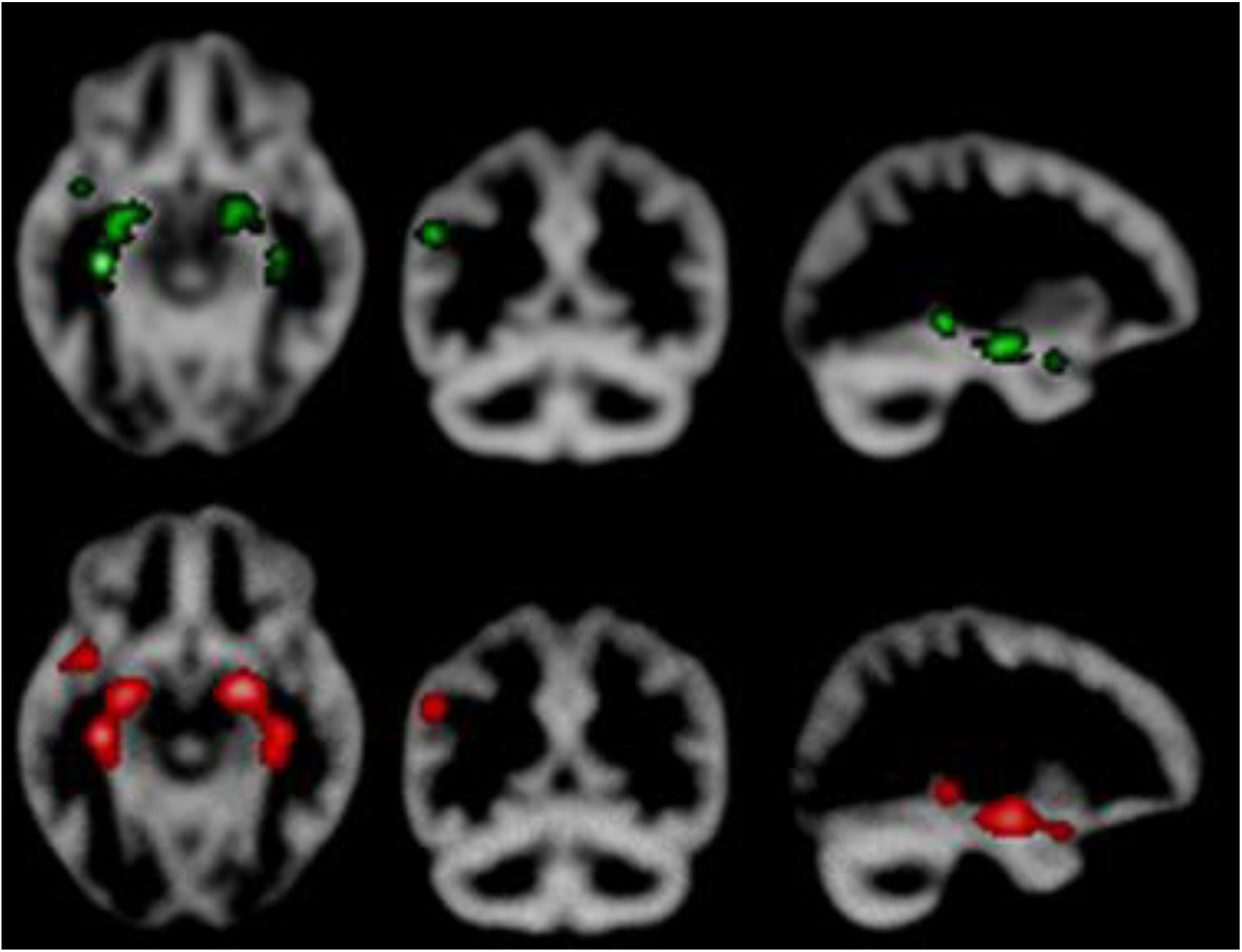
Analysis of structural studies of Alzheimer’s disease. The results from CVCA (top) and ALE (bottom) are similar. In this example the Z score from CVCA null hypothesis significance testing was 62, the kernel width was 5.5mm, and the *N*_*min*_ parameter was 4.

**Figure 6.**
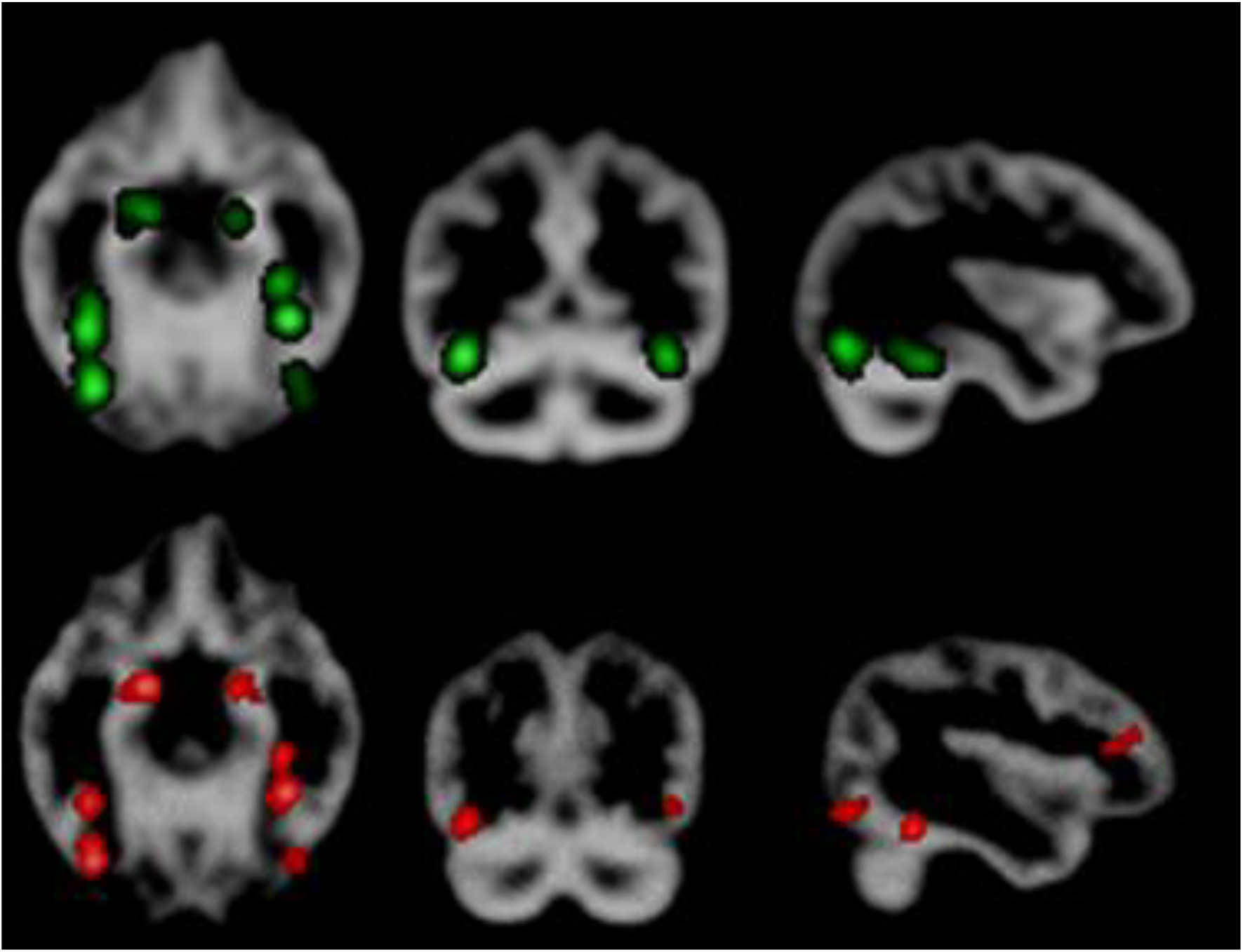
Analysis of functional face discrimination studies. The results from CVCA (top) and ALE (bottom) are similar. In this example the Z score from CVCA null hypothesis significance testing was 48, the kernel width was 8.5mm, and the *N*_*min*_ parameter was 4.

### Narcolepsy

This involved 15 studies of mixed modality. The results (figure 7 (top)) show a uniform distribution of foci. Given the null hypothesis of uniformly distributed foci, it is not surprising that these results are not significant, with an estimated Z score of 0.02; this was also the finding using the ALE algorithm (Rahimi-Jafari et al., 2022).

**Figure 7.**
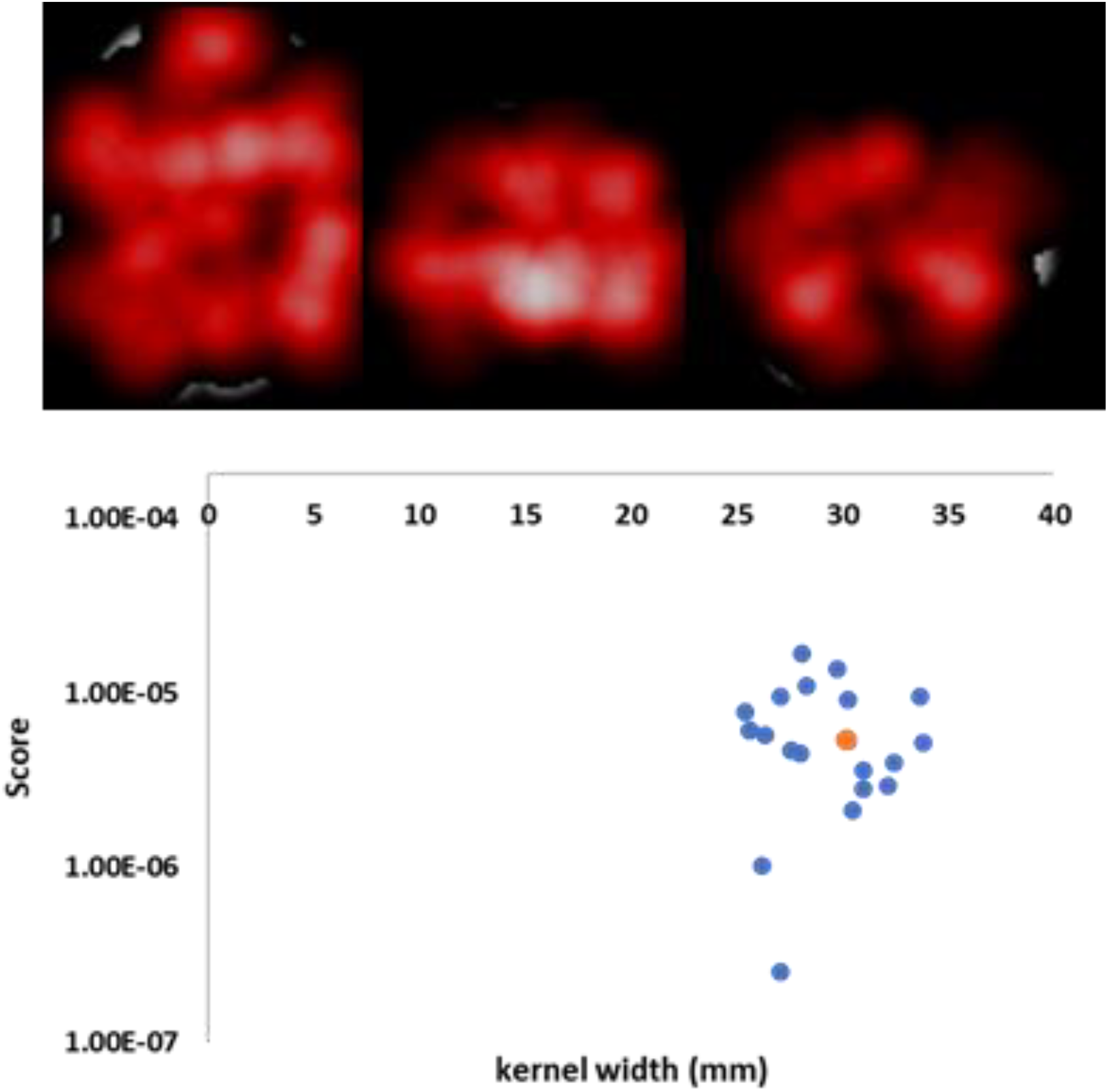
The reported foci are distributed quite uniformly throughout the grey matter, as depicted by the density (top) estimate from CVCA. The score for the reported data falls centrally within the scores of the null data, suggesting the result is not significant; Z score=0.02. The *N*_*min*_ parameter was 5 and the kernel width 30mm.

## Discussion

Here a CBMA algorithm has been presented that does not rely on null hypothesis significance testing to threshold results and does not use an empirically fixed kernel width that might not be valid for all cases. Instead, a cross-validation approach is used in an attempt to estimate the reported foci density by forcing that density to be a maximised near the foci of the left-out study. The analysis may be performed stand-alone, or to verify that meta-analyses are providing similar results to increase confidence overall by the use of cross-validation.

The use of empirical kernel widths in CBMA algorithms means that results may not be valid under some conditions. For example, if the meta-analysis involves very many independent studies, it is likely that a smaller kernel would be appropriate (Epanechnikov, 1969) to indicate how the reported foci in different studies are related. Conversely, a meta-analysis involving few studies would likely require a larger kernel as the mean distance to the nearest foci from the different studies would be larger. Statistical thresholding using NHST is quite normal practice in CBMA, and in neuroimaging in general, and is a pragmatic solution to a problem. However, such thresholding is most interpretable in a prospective setting, where an experiment can be designed to force (with some small type 2 error rate) a correctly hypothesised effect to be significant. In meta-analysis there is no way to modify the sample of data analysed to define the power of the study. Indeed, the typical aim of meta-analysis is not NHST, but better estimation of effect.

It is not feasible to validate every CBMA performed to show that the empirical kernel and NHST are producing reliable results, however it is always possible to use cross-validation. That is the purpose of CVCA. By conducting a meta-analysis on most of the data, the kernel and thresholding can be optimised to reflect the data that are held out. While this is not a complete validation of the results, it does offer some hint at how generally valid the results may be.

Comparison of the results of CVCA to the popular ALE algorithm has been conducted here. The analysis of Alzheimer’s disease voxel-based morphometry studies, and the analysis of face discrimination fMRI studies revealed that ALE and CVCA produced similar results for these examples. Other comparisons have also been conducted (not shown), and also revealed similar results. In the Narcolepsy example, CVCA produces a density estimate that suggests a uniform spread of foci across the brain, which is therefore similar to the null and not significant.

CVCA analyses only the summary results from multiple studies to estimate the density of reported foci relating to a hypothesis. Therefore, there is little spatial data available for a more accurate estimate. For example, with more data the kernel width might be considered different in different parts of the brain, and this could be considered within the optimisation process. Furthermore, while testing different kernel profiles produced largely similar results (not shown), there may be a more appropriate kernel not considered. There are also more options for the score function, for example weighting the contribution by study sample size, but this has not been explored due to lack of gold standard to decide which is superior. Another problem for CVCA is that studies reporting no significant foci (‘null’ studies) do not contribute to the analysis. For these ‘null’ studies it is assumed that the same effect was detected but did not reach statistical significance. When there are many such ‘null’ studies, the results of CVCA should be considered potentially biased.

## Summary

CBMA typically uses a kernel of fixed (against the number of studies) width, which may not be optimal for all cases as kernel density estimation (for example) requires a reducing kernel width with more data. Furthermore, the use of null hypothesis statistical testing to define the results is different to conventional meta-analysis where the aim is to compute better estimates of effect. Unfortunately, it is not feasible to validate these features in every possible case. Here CVCA is introduced, which performs cross-validation for every analysis. CVCA does not have a fixed kernel width and does not use null hypothesis statistical testing to define its results. Instead, results are determined by imposing the constraint that the kernel density estimate of reported foci is maximised at the reported foci of the left-out study. Currently CVCA will produce a reported effect density estimate and a report of brain regions affected. Further analyses are in development.

## Data Availability

All data produced in the present study are available upon reasonable request to the authors

